# The impact of higher uptake of plant-based diets in England: model-based estimates of health care resource use and health-related quality of life

**DOI:** 10.1101/2023.12.26.23300536

**Authors:** Nadine Henderson, Chris Sampson

## Abstract

Plant-based diets have grown in popularity in recent years, in part because of the perceived health benefits; higher consumption of vegetables and other healthy foods is associated with better nutrition and reduced risk of disease. In this study, we estimate the potential impact of higher uptake of a 100% plant-based (vegan) diet in England from the perspective of the National Health Service (NHS). We estimate the impact in terms of quality-adjusted life years (QALYs), health care expenditure, and total net benefit compared to the current level of uptake.

This model-based analysis combines estimates for disease prevalence, the relative risk of disease associated with a vegan diet, and disease-specific health state utility values (HSUVs) and health care costs. We conducted a literature review to identify the most suitable inputs to the model, which included estimates for cancer, coronary heart disease, cataracts, diverticular disease, bone fractures, stroke, and type 2 diabetes. The model is open-source and implemented in an interactive online dashboard, allowing for further extension and exploration of the findings.

In our base case analysis with 100% adoption of a plant-based diet in England, the total health care cost savings across all considered diseases is around £6.7 billion per year, with 172,735 additional QALYs, and a total net benefit to the NHS of around £18.8 billion when QALYs are valued at £70,000. The majority of potential savings are realised through the avoidance of almost 1.3 million cases of type 2 diabetes.

Numerous challenges are associated with estimating the impact of widespread dietary changes in society. However, strong evidence shows that plant-based diets are associated with better health outcomes for some of England’s most significant causes of disease burden. Higher rates of plant-based diet adoption may bring considerable cost savings for the NHS and generate substantial health benefits for the population. Policymakers should consider the relevance of these estimates to their settings and the potential for interventions that support healthy dietary changes that contribute to population health. Future research should seek to identify the causal effects of plant-based diet adoption on health outcomes, and health care resource use across different populations.

**Funding:** This study was funded by a grant from The Vegan Society.

**Author Declarations:** The authors are employees of the Office of Health Economics, a registered charity and independent research organisation that receives funding from a variety of sources. Both authors identify as vegan. CS is a member of The Vegan Society.

## Introduction

The uptake of vegan and other plant-based diets has grown in popularity in the 21st century. People cite various reasons for adopting a plant-based diet, including perceived health benefits, reduced environmental impact, and improved animal welfare (1). Recent statistics suggest that the number of vegans in the UK has more than quadrupled in the last eight years and that vegans currently account for around 3% of the UK population (2,3).

The term ‘plant-based diet’ may be used to refer to a range of distinct diets, from omnivorous diets with low animal source content (e.g., meat and fish) and vegetarian diets (lacto-ovo-vegetarian: plant-based except for dairy products and eggs) to vegan diets (100% plant-based). Plant-based diets may include higher consumption of healthy foods such as vegetables, fruits, nuts, and whole grains (4), associated with higher dietary fibre and less saturated fat (5), which may give rise to differences in health outcomes. The British Dietetic Association states that carefully planned plant-based diets can support healthy living at every age and life stage (6), and the National Health Service (NHS) offers similar guidance for eating a healthy plant-based diet (7).

A growing body of research suggests that plant-based diets may improve health outcomes relating to the most significant causes of disease burden in advanced economies, including coronary heart disease (8) and cancer (9,10). However, relatively little research has estimated the impact of a plant-based diet on health care resource use, or the value of plant-based diets from a health service perspective. One exception is a study by Lin et al., which found that medical expenditure was around 15% lower for vegetarians compared with omnivores in Taiwan (11). This finding was used to approximate potential savings in the UK of around £30 billion per year (12).

Given the expected health benefits associated with plant-based diets, a higher uptake has the potential to improve the health of the population of England and generate considerable cost savings for the NHS. This study seeks to estimate the expected impact associated with a greater proportion of the English population following a plant-based diet. We focus on a vegan diet, which is 100% plant-based and most clearly defined and distinguishable from an omnivorous diet.

Our study focuses on outcomes that are of interest to the NHS and to decision-making at a high level. We estimate expected differences in the total number of cases of relevant diseases, differences in the total number of quality-adjusted life years (QALYs) attained by the population, and differences in total NHS expenditure for one year.

## Methods

This study aims to identify the differences in health care costs and QALYs associated with a society in which a greater proportion of people have adopted a plant-based diet. We developed a model to estimate these outputs using the most relevant available inputs from published literature. The overall approach combines estimates of the relative risk of disease associated with a vegan diet with estimates of the prevalence, costs, and QALYs associated with specific diseases.

### Literature Review

We reviewed published literature on the health impact of vegan diets through a targeted literature search of PubMed and Google Scholar. We initially sought to identify systematic reviews of the health effects of plant-based diets. We then supplemented this with papers relating to the costs or QALYs associated with plant-based diets and diseases identified as being influenced by a plant-based diet.

The main aim was to identify health conditions for which a statistically significant difference in the relative risk (positive or negative) of experiencing the health condition existed for vegan compared with non-vegan diets, preferably derived from a meta-analysis. For each condition identified, we extracted quantitative estimates relating to the i) prevalence, ii) health care costs, and iii) health state utility value (HSUV) associated with the condition. We selected the most relevant estimates for the population of England. Terms used as part of the search strategy are provided in Box 1.

#### BOX 1

LITERATURE SEARCH TERMS

Preliminary search

- systematic review”[Publication Type] AND (“diet, vegan”[MeSH Terms] OR (“diet”[All Fields] AND “vegan”[All Fields]) OR “vegan diet”[All Fields] OR “veganism”[All Fields] OR “vegans”[MeSH Terms] OR “vegans”[All Fields] OR “vegan”[All Fields])
- (“economical”[All Fields] OR “economics”[MeSH Terms] OR “economics”[All Fields] OR “economic”[All Fields] OR “economically”[All Fields] OR “economics”[MeSH Subheading] OR “economization”[All Fields] OR “economize”[All Fields] OR “economized”[All Fields] OR “economizes”[All Fields] OR “economizing”[All Fields]) AND (“diet, vegan”[MeSH Terms] OR (“diet”[All Fields] AND “vegan”[All Fields]) OR “vegan diet”[All Fields] OR “veganism”[All Fields] OR “vegans”[MeSH Terms] OR “vegans”[All Fields] OR “vegan”[All Fields])
- (“quality adjusted life years”[MeSH Terms] OR (“quality adjusted”[All Fields] AND “life”[All Fields] AND “years”[All Fields]) OR “quality adjusted life years”[All Fields] OR “qaly”[All Fields]) AND (“diet, vegan”[MeSH Terms] OR (“diet”[All Fields] AND “vegan”[All Fields]) OR “vegan diet”[All Fields] OR “veganism”[All Fields] OR “vegans”[MeSH Terms] OR “vegans”[All Fields] OR “vegan”[All Fields])

Condition-specific searches

- “Disease/condition” AND “prevalence” OR “incidence” OR “total cases”
- “Disease/condition” AND “Economic burden” OR “NHS expenditure” OR “NHS spending” OR “Cost”
- “Disease/condition” AND “Health state utility” OR “Health state value” OR “EQ-5D utility”

### Modelling Tool

We developed a modelling tool to estimate changes in i) total cases, ii) QALYs, and ii) total health care costs. The model uses parameters extracted from the literature review. We developed the model in R and created an interactive web application using Shiny (13), which allows for manipulating all model inputs and live reading of corresponding estimates^1^. The structure of the model is shown in Figure 1, with model inputs shown in grey boxes, outputs in green boxes, and arrows showing which inputs were used to calculate each output.

**FIGURE 1:**
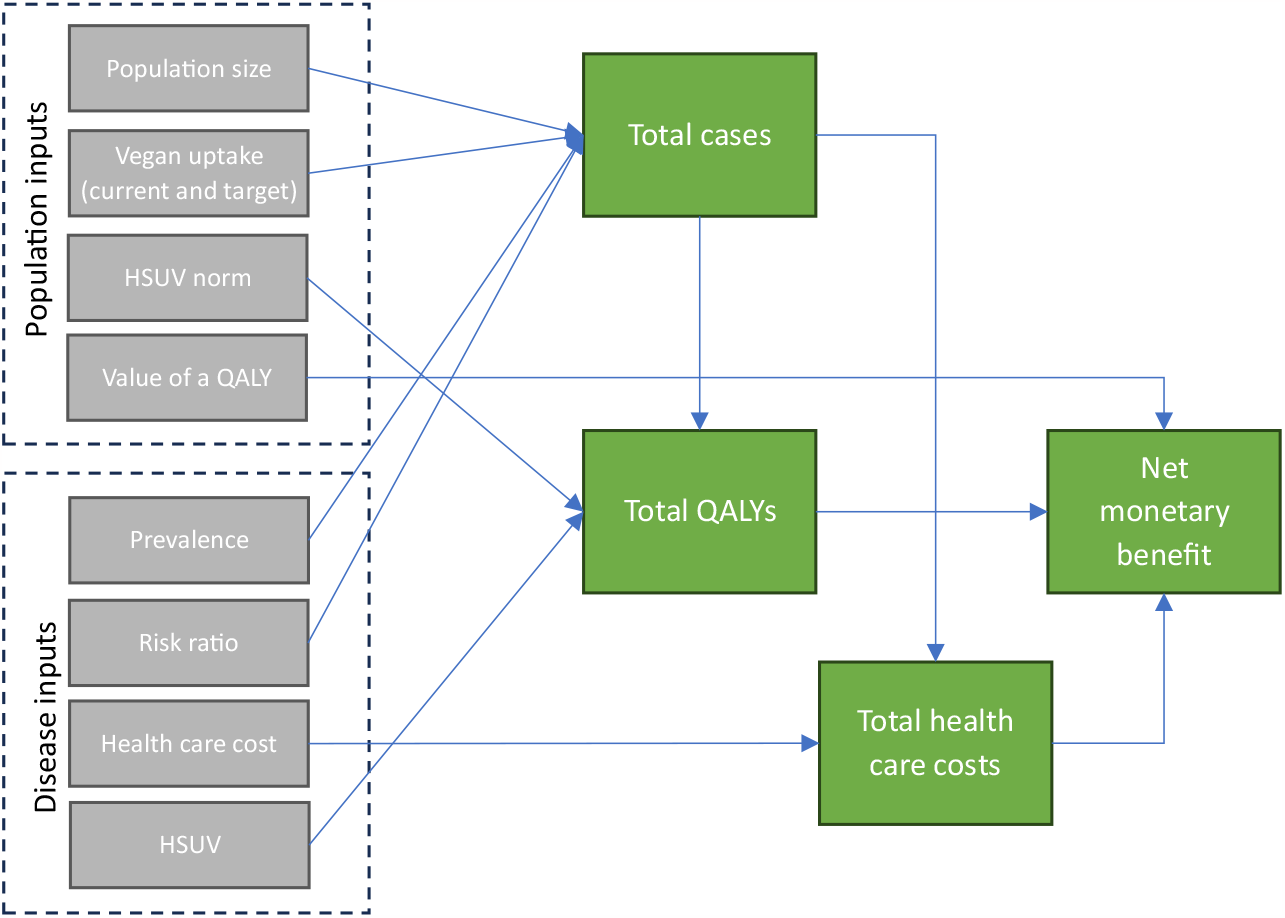
MODEL STRUCTURE.

### Base case analysis

The flexible modelling tool facilitates an unlimited range of scenario analyses based on our specified inputs, including the option to introduce an additional health condition. In this paper, we focus on a base case analysis for the population of England, assuming that 3% of the population is currently vegan and with an uptake of 100% for comparison. The base case analysis defines the default values within the modelling tool.

We used recent population estimates for England reported by the Office of National Statistics (14). Disease-specific HSUVs were subtracted from a population norm HSUV, representative of the general population, to estimate the QALY losses associated with disease. For the base case, we used an HSUV population norm based on UK EQ-5D time trade-off (TTO) values reported by Janssen and Szende (15).

We extracted condition-specific HSUVs from a catalogue of EQ-5D-3L values for chronic conditions and health risks (16). If the HSUV for a condition was unavailable in the catalogue, we identified applicable EQ-5D values from peer-reviewed articles, with a preference for systematic reviews.

Results are presented in an impact inventory to report the outcomes associated with a vegan diet in England in a disaggregated way. The impact inventory summarises the outputs of the economic model, for each disease and overall, including:

i. Difference in the number of cases
ii. Difference in QALYs
iii. Difference in health care costs

We also estimated the net monetary benefit associated with the level of vegan diet uptake by multiplying the number of QALYs by a monetary value of £70,000, derived from the UK HM Treasury Green Book (17), and adding this to the health care cost savings.

All base case model inputs are described in Table 1.

**TABLE 1:**
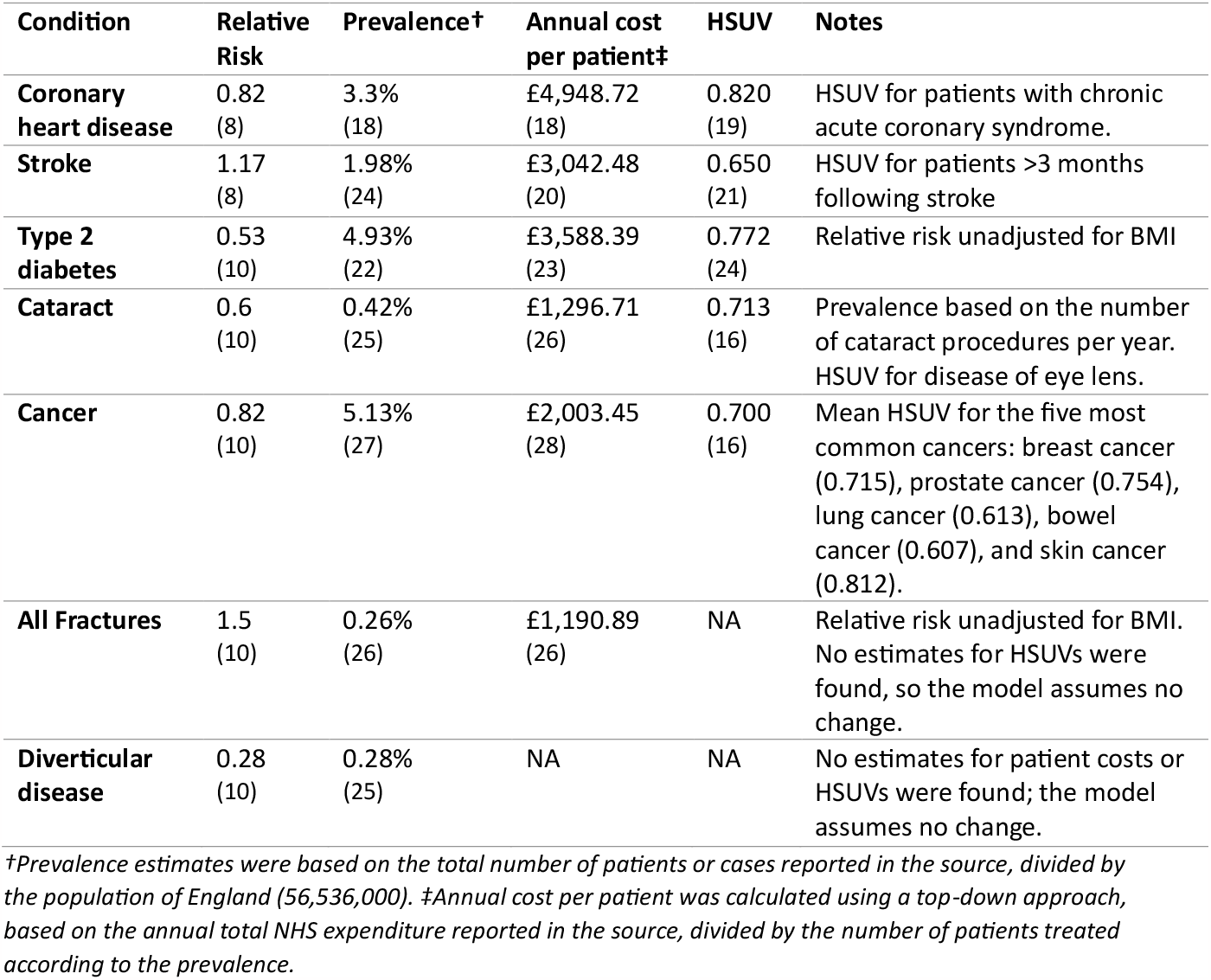
BASE CASE MODEL INPUTS AND SOURCES.

### Scenario analyses

In addition, we report on some alternative scenarios. Scenario 1 assumes an additional 10% adoption of a vegan diet in England, from 3% to 13% of the population. Scenario 2 uses BMI-adjusted relative risk estimates where available.

## Results

### Literature review

We identified a decreased relative risk associated with a plant-based diet for cancer (10), cardiovascular disease (8,29,30), cataracts (10), ischaemic heart disease (8,9), type 2 diabetes (10), diverticular disease (10), and obesity (31). We identified an increased relative risk associated with a vegan diet for stroke (8,10) and fractures (10).

The decreased risk of cancer was estimated for all cancers combined, which was 18% lower for vegans compared to meat-eaters (10). There is variation in the direction of risk for different cancer sites. For example, estimates suggest that vegans have a lower risk of stomach and haematological cancers but have a higher risk of cervical cancer, all with wide confidence intervals due to the small sample sizes used in the research.

Cardiovascular disease had the most extensive evidence indicating a lower risk associated with plant-based diets (8,29,30). A decreased risk of coronary heart disease (referred to as ischaemic heart disease) was reported by Dybvik et al. (8) and Dinu et al. (9). A recent meta-analysis found that plant-based diets were associated with lower levels of total cholesterol and low-density lipoprotein cholesterol, which are risk factors for cardiovascular diseases (32).

A systematic review found that the hazard ratio of stroke for those following a vegan diet compared to meat-eaters was 1.17 (95% CI: 0.69-1.99), though this was not statistically significant (8). Key et al. (10) found a hazard ratio of stroke for vegetarians of 1.17 (95% CI: 1.00-1.38) but had insufficient data to estimate the risk for vegans.

Key et al. (10) found that the risk of diabetes in vegans was 47% lower than in meat-eaters. This was reduced to 1% after adjusting for BMI, implying that lower BMI explains the lower risk in vegans. In the economic model, we use the BMI unadjusted risk of diabetes to account for the reduced weight and lower BMI of vegans compared to meat-eaters, which are both risk factors associated with type 2 diabetes (33).

There is good evidence that plant-based diets are associated with a reduced risk of obesity. For example, Chiu et al. (31) found that each additional year of a vegan diet lowered the risk of obesity by 7% for Taiwanese adults, and similar results have been reported for the UK (34). However, our base case analysis excludes obesity from the model because we expect obesity to be a key risk factor overlapping with other disease outcomes captured elsewhere. This is especially true for type 2 diabetes, where it is likely that we would be double-counting any benefits associated with a plant-based diet.

The BMI-adjusted risk of bone fractures is higher among vegans (43%) and vegetarians (9%) compared to meat-eaters (50% and 11% unadjusted for BMI) (10). This interaction with BMI is notable; in vegans, the relative risk for hip fracture was 3.17 among people with a BMI of less than 22.5 kg/m2 but 0.94 for people with a BMI of 22.5 kg/m2 and above. However, the authors note that this finding was based on small numbers in the subgroups. A vegan diet may be associated with relatively lower intakes of calcium and vitamin D (10), and lower bone mineral density has been observed in people with a vegan diet (35), though the methods underlying this finding have been challenged (36).

A systematic review of plant-based diets compared with mental health outcomes found no significant association between diet and depression scores, stress, or well-being (37).

In a study of vegetarian diets and mortality in a cohort of over 70,000 Seventh-Day Adventists, the adjusted hazard ratio for all-cause mortality in vegans was 0.85, and in lacto-ovo-vegetarians it was 0.91 (38). Significant associations with vegetarian diets were detected for cardiovascular mortality, non-cardiovascular non-cancer mortality, renal mortality, and endocrine mortality (38). In our analysis, we conservatively exclude mortality outcomes. Our analysis assumes that impacts are realised over a year and does not facilitate observation of events at different times.

The sources used in our study for the relative risk of disease compared omnivorous diets with plant-based diets that excluded all animal products, including dairy and eggs (8,10). In these studies, most of the dietary information was derived from dietary and food frequency questionnaires. The reported risk estimates were based on prospective cohort studies.

### Base case analysis

In our base case analysis with 100% adoption of a vegan diet in England, with a population of 56,536,000, the total health care cost savings across all considered diseases is estimated to be around £6.7 billion. Results are summarised in Table 2.

**TABLE 2:**
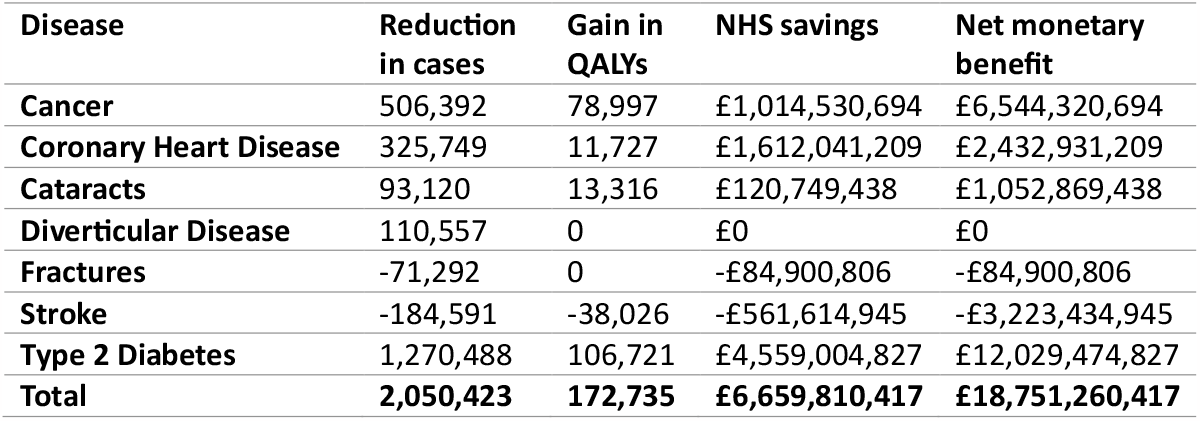
BASE CASE ANALYSIS (100% UPTAKE)

| Disease | Reduction in cases | Gain in QALYs | NHS savings | Net monetary benefit |
| --- | --- | --- | --- | --- |
| **Cancer** | 506,392 | 78,997 | £1,014,530,694 | £6,544,320,694 |
| **Coronary Heart Disease** | 325,749 | 11,727 | £1,612,041,209 | £2,432,931,209 |
| **Cataracts** | 93,120 | 13,316 | £120,749,438 | £1,052,869,438 |
| **Diverticular Disease** | 110,557 | 0 | £0 | £0 |
| **Fractures** | -71,292 | 0 | -£84,900,806 | -£84,900,806 |
| **Stroke** | -184,591 | -38,026 | -£561,614,945 | -£3,223,434,945 |
| **Type 2 Diabetes** | 1,270,488 | 106,721 | £4,559,004,827 | £12,029,474,827 |
| **Total** | **2,050,423** | **172,735** | **£6,659,810,417** | **£18,751,260,417** |

For a 100% uptake of a vegan diet, the total QALYs gained would be 172,735, with a value of around £12.1 billion when QALYs are valued at £70,000. The expected net monetary benefit to the NHS of resource use savings and QALY gains combined is £18.8 billion per year if 100% of the population adopts a vegan diet.

These results are primarily driven by impacts on type 2 diabetes, for which we estimate 1.3 million fewer cases, savings of £4.6 billion, and 106,721 additional QALYs per year.

### Scenario analyses

In our first scenario analysis, we assume that an additional 10% of the population of England adopts a vegan diet (i.e., 13%), which is likely to be a more realistic target. The estimated total health care cost savings across all considered diseases is around £687 million. For an additional 10% uptake of a vegan diet, the total QALYs gained would be 17,808, with a value of around £1.2 billion. The expected net monetary benefit to the NHS of resource use savings and QALY gains is £1.9 billion per year if an additional 10% of the population adopts a vegan diet. Results are summarised in Table 3.

**TABLE 3:** SCENARIO ANALYSIS (10% ADDITIONAL UPTAKE)

| Disease | Reduction in cases | Gain in QALYs | NHS savings | Net monetary benefit |
| --- | --- | --- | --- | --- |
| **Cancer** | 52,205 | 8,144 | 104,590,793 | £674,670,793 |
| **Coronary Heart Disease** | 33,582 | 1,209 | 166,189,815 | £248,299,209 |
| **Cataracts** | 9,600 | 1,373 | 12,448,396 | £108,558,396 |
| **Diverticular Disease** | 11,398 | 0 | £0 | £0 |
| **Fractures** | -7,350 | 0 | -8,752,660 | -£8,752,660 |
| **Stroke** | -19,030 | -3,920 | -57,898,448 | -£332,298,448 |
| **Type 2 Diabetes** | 130,978 | 11,002 | 470,000,498 | £1,240,140,498 |
| **Total** | **211,383** | **17,808** | **686,578,394** | **£1,930,617,788** |

In a second scenario analysis, we assume that 100% of the population of England adopts a vegan diet, but we adjust the model’s relative risk inputs for BMI where possible. BMI-adjusted estimates were available for fractures and type 2 diabetes. In this case, the total health care cost savings across all considered diseases is estimated to be around £2.2 billion. The total QALYs gained would be 68,285, valued at around £4.8 billion. The expected net monetary benefit to the NHS of resource use savings and QALY gains is therefore £7 billion per year if 100% of the population adopted a vegan diet. After adjusting for BMI, the results are primarily driven by impacts on cancer, for which we estimate over 500,000 fewer cases, savings of £1 billion, and almost 80,000 QALYs. Results are summarised in Table 4.

**TABLE 4:** SCENARIO ANALYSIS (100% UPTAKE AND BMI-ADJUSTED RISK)

| **Disease** | **Reduction in cases** | **Gain in QALYs** | **NHS savings** | **Net monetary benefit** |
| --- | --- | --- | --- | --- |
| **Cancer** | 506,392 | 78,997 | £1,014,530,694 | £6,544,320,694 |
| **Coronary Heart Disease** | 325,749 | 11,727 | £1,612,041,209 | £2,432,931,209 |
| **Cataracts** | 93,120 | 13,316 | £120,749,438 | £1,052,869,438 |
| **Diverticular Disease** | 110,557 | 0 | £0 | £0 |
| **Fractures** | -61,311 | 0 | -£73,014,693 | -£73,014,693 |
| **Stroke** | -184,591 | -38,026 | -£561,614,945 | -£3,223,434,945 |
| **Type 2 Diabetes** | 27,032 | 2,271 | £97,000,103 | £255,970,103 |
| **Total** | **816,948** | **68,285** | **£2,209,691,806** | **£6,989,641,806** |

## Discussion

### Main findings

We have estimated significant value to the NHS associated with the wider adoption of a plant-based diet in England. The differences in outcomes resulted from an expected reduction in cases of cancer, coronary heart disease, cataracts, and diabetes.

Research on the health care resource impact of a vegan diet is limited, and there are few studies with which to compare our results. The most similar research we identified was by Lin et al. (11), who found that vegetarians have a lower rate of outpatient visits to the doctor because of their healthier diets, which tend to be high in fruit, vegetables, and whole grains. This translated into a 15% lower total medical expenditure per person compared with people who eat meat, particularly for chronic illnesses such as high blood pressure, heart disease, and depression. Following the publication of this research, a 2022 news article cited Dr Shireen Kassam, who founded the Plant Based Health Professionals network (12). Dr Kassam calculated that if a similar 15% decrease was observed in the UK, the NHS could save £30 billion annually. This estimate was based on health care expenditure in 2019 of £225.2 billion, equating to £3,371 per person.

The quoted £30 billion estimate is significantly higher than our estimate of £6.7 billion. There are numerous possible reasons for this, and our assessment relies on considerably more inputs and more transparent assumptions. Our estimate relates to England, rather than the UK, and we do not include all causes of disease or potential health impacts of a plant-based diet, as we discuss in detail below. Furthermore, our analysis employs several conservative assumptions. For instance, we exclude the possible implications of obesity as distinct from other health problems, and we assume there to be no impact on costs or QALYs where evidence is unavailable (i.e., for diverticular disease and fractures).

### Limitations

There are numerous limitations to our study. Many limitations are unavoidable for an ambitious estimation strategy based on limited evidence. The transparency of our analysis, with an interactive web application and public release of the underlying code, ensures that researchers and other stakeholders can assess the importance of these limitations.

### Evidence availability

Most of the evidence available for the relationship between plant-based diets and health outcomes is based on non-experimental observational studies. These studies observe people who have chosen different dietary habits and identify correlations between dietary patterns and health outcomes. Therefore, they can only tell us about the health effects of a vegan diet *for people who choose to adopt a vegan diet*. Whilst confounding factors are usually accounted for, there are likely to be underlying factors not included, which may affect estimates. In particular, there is likely to be a significant selection issue in these studies, whereby the people who choose to adopt a plant-based diet are different to those who don’t in important ways. The implications of this for our research are unknown; the impact of a plant-based diet for people who do not choose to adopt a plant-based diet may be greater or lesser than our findings suggest. However, where experimental evidence relating to plant-based diets is available, it tends to concur with the results of non-interventional studies (39).

Due to the limited evidence base, we needed to make numerous assumptions when selecting parameters. For instance, we used HSUVs – including a population norm HSUV – that do not correspond to groups adopting particular diets. In some cases, we also averaged different HSUVs to identify a suitable input for our model. We also made significant assumptions about the estimated risk of disease. For instance, our model adopted hazard ratio and relative risk estimates and assumed them to be equivalent to the relative risk of disease in the population for the one-year time horizon of our model.

### Our modelling approach

Our model aims to describe a world in which a greater proportion of the population adopts a vegan diet, thus capturing the preventative potential of vegan diets. A model of this kind necessarily involves many assumptions and simplifications.

Our model adopts a 1-year time horizon, assuming that the prevention value associated with a plant-based diet is realised within one year for all conditions. The effects of a plant-based diet are likely to develop over time, and our model is therefore not suited to estimating the short-term benefits of adopting a plant-based diet. This particular assumption is expected to overestimate differences in the short term.

Another limitation of our modelling approach is that it cannot capture the complete range of ways in which dietary changes might impact health outcomes. Our model attempts to capture the preventative impact of a vegan diet on various health conditions, which may be the most important mechanism by which a plant-based diet can improve health. However, evidence suggests that a plant-based diet could be beneficial in managing certain chronic diseases, such as type 2 diabetes (40), which may be driven by lower body weight and general diet quality. Our model will likely produce underestimates because it only accounts for value in primary prevention.

Furthermore, we have focused exclusively on realised health outcomes, while a significant amount of research relates to intermediate outcomes and risk factors associated with diet and disease. For example, evidence suggests that people following a vegan diet have lower weight, lower BMI, lower cholesterol, and lower blood pressure. These factors are associated with many conditions, including diabetes and the other diseases in our model.

The most critical set of risk factors related to different diets and diseases is, arguably, nutrient intake. In a systematic review of nutrient intakes of vegan, vegetarian, and omnivorous diets across Europe, South/East Asia, and North America, Neufingerl and Eilander (41) observed nutrient deficiencies across all dietary patterns, highlighting the importance of a well-planned diet regardless of dietary preference, and our model cannot capture these dynamics. Nutritional intake is a critical factor in determining health outcomes and is not captured in our model. The diets consumed by participants of the included studies (both omnivorous and vegan) are likely to reflect the eating habits of the general population, with variation in how healthy the studied diets are.

Our model is also limited in its capacity to explore different realistic scenarios. With additional evidence and a more sophisticated model, sex- and age-specific population norms could be used to generate more accurate estimates of QALYs generated. In addition, mortality estimates could have been incorporated to fully capture the potential increase in length of life resulting from avoiding some acute and severe conditions.

### Exclusion of relevant health outcomes

Many potentially relevant health outcomes are not included in our model. Most notably, our model does not fully capture the impact of lower weight and BMI associated with a vegan diet, and the implications that this would have for people who are overweight or obese. Obesity is a risk factor associated with many acute and chronic conditions, which could not all be included in this study. Health care interventions such as medication regimes and surgery may also be more effective in people with a healthy weight. Furthermore, more transitions to a plant-based diet could decrease the number of obesity-related interventions such as bariatric surgery. Conversely, some people may be at greater risk of being underweight with a vegan diet. We also exclude mortality from our model, though we would expect its inclusion to increase the magnitude of our findings.

### Exclusion of non-health outcomes

In this study, we have estimated health effects and direct health system costs. However, there is likely to be a net positive impact on informal health care, social services, productivity, and the environment. These may benefit the overall economy, health service, and public well-being. Dramatic shifts in dietary patterns have the capacity to bring significant improvements across every aspect of individuals’ lives, all areas of public policy, and planetary health, and we have not sought to capture all of these in our study.

### Implications

Our study suggests that a greater uptake of a plant-based diet could have considerable net health benefits and cost savings for the NHS. Public health decision-makers should consider strategies to support people in transitioning to more healthy diets, including vegan diets.

The limited availability of data and clinical research highlights substantial gaps in our understanding of plant-based diets’ potential health and broader benefits. Few publicly available or otherwise large datasets describe people’s dietary patterns alongside health outcomes. One important exception is the European Prospective Investigation into Cancer and Nutrition (EPIC) study, but even this has limited information on generic health outcomes and health care resource use. Our study signals the need for new data collection initiatives, reporting clinical and health outcomes for people following different dietary patterns, including vegan diets.

Further research is needed to understand and validate the impact of a plant-based diet across health conditions, including site-specific cancer types, which have not been investigated thoroughly. Prospective and interventional research should be a priority to support causal inference for the impacts of adopting a plant-based diet.

## Conclusion

Higher rates of plant-based diet adoption in England could bring significant benefits to the NHS and population health. If everybody in England were vegan, NHS expenditure could be reduced by £6.7 billion per year, with 2 million fewer cases of disease and a gain of more than 170,000 quality-adjusted life years across the population. The total value to the NHS could be around £18 billion per year. These findings rely on a limited evidence base, and future research should seek to identify robust estimates for the impact of a plant-based diet on health-related outcomes.

## Data Availability

All data produced in the present work are contained in the manuscript.

https://chrissampson87.shinyapps.io/VeganDiet/

## Footnotes

1 The app can be accessed at https://chrissampson87.shinyapps.io/VeganDiet/

